# High Schistosomiasis Prevalence in Urban and Rural Primary School Children from Kimpese Health District, Democratic Republic of the Congo

**DOI:** 10.64898/2026.09.24.26363880

**Authors:** Kevin Lubula Karume, Harry Noyes, Oscar Nyangiri, Julius Mulindwa, Olivier Fataki, Oscar Kwabanzawoko, Ronald Futila Kyong Shin, Jacent Nassuuna, Allison Elliott, Miriam Casacuberta-Partal, Jacob Lyaki, Jean Pierre Kambala Mukendi, Theophile Fund Mbemba, Dieudonne Mumba Ngoyi, the TrypanoGEN+ research group of the H3Africa consortium

## Abstract

**Background:** Schistosomiasis is a parasitic disease caused by flatworms known as schistosomes. The infection cycle starts when people shed eggs through the stool or the urine. Larva infects freshwater snails to later infect humans through the skin during water based activities. Schistosomiasis is commonly associated with rural areas. In this study, we wanted to see if the schistosomiasis distribution is different in rural and urban areas, along with potential risk factors associated with schistosomiasis distribution.

**Methodology:** Fresh urine samples were collected from 1,699 school-aged children (5 – 15 years old) in urban and rural sites in the Kimpese health district. Prevalence of haematobium and mansoni schistosomiasis was determined using, respectively, the urine sedimentation and the Point-of-Care Circulating Cathodic Antigen test with a quantitative estimate of infection intensity obtained with an RDT-Reader. Collected risk factors were tested for association with schistosomiasis infection and its intensity.

**Results:** The overall prevalence of schistosomiasis found in urban and rural areas was 75% and 82% respectively, with higher prevalence of *S. mansoni* in rural schools, and higher *S. haematobium* prevalence in urban schools. The difference in total prevalence between schools in urban and rural areas was significant, X^2^ (1, N = 1699) = 13.03, p = .0003. *S. mansoni* intensity of infection was also higher in rural children (mean = 162 mV) compared to those in urban areas (mean = 76 mV) (p < 2e-16).

Male gender, washing dishes in the river, and swimming were associated with schistosomiasis in urban areas. Age, Springs, and rivers were significant factors in rural sites. Age, river, and washing dishes in the river were factors associated with the intensity of infection.

**Conclusion:** Our results showed very high prevalence in both rural and urban areas, which implies that control programs should consider them with the same interest and also revise control strategies.

**Author Summary:** Schistosomiasis is a common but often overlooked parasitic disease that affects millions of people around the world. More than 290 million people are at risk of becoming infected. The disease is mainly spread when people come into contact with freshwater sources, such as rivers and streams, that contain infected snails carrying the parasite.

In this study, we tested 1,699 schoolchildren aged 5 to 15 years living in both urban and rural communities of the Kimpese district in Kongo Central Province, Democratic Republic of the Congo. Using a rapid diagnostic test (POC-CCA), we found that schistosomiasis was very common in both settings, affecting about 75% of children in urban areas and 82% in rural areas.

Intestinal schistosomiasis was more common in rural communities, while urinary schistosomiasis was found more frequently in urban communities. Factors such as being male, age, and activities involving river water, especially swimming and washing dishes, were linked to a higher risk of infection.

These findings show that schistosomiasis is not only a rural health problem. Even in urban areas where access to safer water may be better, people remain at risk if they continue to have regular contact with rivers and other freshwater sources that contain the parasite.

## Introduction

Schistosomiasis is a disease caused by trematode parasites of the genus *Schistosoma,* which use freshwater snails as intermediate hosts. *Schistosoma mansoni* and *Schistosoma haematobium* are the main human-infecting species found in Africa, causing respectively intestinal and urogenital schistosomiasis in their adult stage in humans ^1^. An estimated 85% of the world’s cases of schistosomiasis occur on the African continent, with over 50% prevalence in some populations ^2^.

Schistosomiasis is traditionally a rural disease of low- and medium-income countries. It is prevalent in populations that use contaminated water for daily activities. Mass drug administration (MDA) has usually focused on rural areas because of the higher prevalence and concentration of cases in such areas. However, many cases have also been reported in urban and peri-urban areas, mainly in Asia and Africa ^3^.

In the Democratic Republic of the Congo (DRC), schistosomiasis is a country-wide infectious disease. 66 surveys have reported it since 1954 in the East, the South, and the West. Over 380 sites have been reported from 1955 to 2015, mostly in rural areas, with the highest prevalence (>60%) for mansoni schistosomiasis recorded in Maniema, Kasai, and Kongo Central ^4^.

Two large surveys on schistosome infection conducted in two rural endemic foci of Kwilu district (Kongo Central) and Kasansa (Kasai), reported a prevalence of 55.8% and 82.7%, respectively, in 5-15-year-old children. They have provided important data on the schistosomiasis epidemiological situation ^5,6^, and those survey only relies on kato-katz However, data on schistosomiasis prevalence in urban and peri-urban areas of Kongo Central remain scarce.

The Point-of-Care Circulating Cathodic Antigen (POC-CCA) assay is primarily used for the diagnosis of Schistosoma mansoni infection because the test was specifically developed and extensively validated for intestinal schistosomiasis caused by this species. The assay detects circulating cathodic antigens released by living adult worms into the bloodstream and subsequently excreted in urine, and *S. mansoni* infections generally produce antigen concentrations that are sufficiently high and consistent for reliable detection ^7,8^. In contrast, the sensitivity of POC-CCA is lower and more variable for other schistosome species, particularly Schistosoma haematobium, making its interpretation less reliable outside *S. mansoni*-endemic settings. Furthermore, several studies and WHO-supported control programs have demonstrated that POC-CCA has higher sensitivity than the traditional Kato-Katz technique method, especially in low-intensity infections where egg excretion is inconsistent ^9–11^. Because Kato-Katz relies on microscopic detection of parasite eggs in stool, it may underestimate prevalence due to day-to-day variation in egg output and reduced sensitivity in light infections. POC-CCA, however, detects active worm antigens independently of egg excretion, allowing it to identify infections that may be missed by stool microscopy. Therefore, in endemic settings and epidemiological surveys targeting *S. mansoni*, the use of POC-CCA alone is considered scientifically valid and operationally advantageous for mapping and surveillance purposes ^12–14^.

In this study, we mapped the prevalence of urinary and intestinal schistosomiasis in several health areas of the Kimpese health district, compared the distribution of schistosome infection in urban and rural area types within that same district, and identified factors associated with their distribution.

## Methods

### Study area

Kimpese is a 12.6 km^2^ town on the main National Highway Number 1, 195 km Southwest of Kinshasa, with a population estimated at over 100,000 people in 2020 ^15^. It is part of Kongo Central Province.

Kongo Central is one of the 26 provinces of DRC located in the western part of the country, with an estimated population of 6,838,500 (2020) in an area of 53,920 km^2^.

Kimpese health district consists of a main urban area (Kimpese City), itself divided into four subdistricts, and a rural area consisting of surrounding villages. The key differences between urban and rural areas are population density, access to clean water, presence of buildings and roads, easy access to high-level health care, and education. Three rural health areas (Malanga, Kiasungua, and Kilueka) and 2 urban health areas, the Centre Evangelique de Cooperation (C.E.Co.) and Kimbanguiste, were selected for surveys on schistosome infection.

### Population, Study design, and Data collection

A school-based cross-sectional study was carried out from the 28^th^ July to the 24^th^ August 2021 in Kimpese City and surrounding villages. Schools were selected randomly in the city of Kimpese, while in rural villages all existing schools were included. In each school all children aged 5-15 years who were willing to provide a urine sample were included in the study, unless they had been treated for schistosomiasis in the past 6 months.

Sociodemographic data, including age, sex, source of water of the household (spring or river), river-related activities such as swimming, washing dishes, or laundry, and the distance between the schools and rivers, were selected as risk factors. Children’s age was categorised into 5 – 10 years old and 11 – 15 years old.

### Detection of schistosome infection

The samples were collected in the morning between 8 a.m. and 12 p.m. About 20 mL of fresh urine was collected, of which 10 mL was centrifuged at 3000 rpm for the detection of *S. haematobium* eggs in the sediment by one microscopist ^16^. Urine samples that were negative for *S. haematobium* were tested for *S. mansoni* using the Point-of-Care Circulating Cathodic Antigen (POC-CCA) test (Rapid Diagnostics South Africa / Batch No. 210412036), and quantified with a Cassette Reader (REF ESLR11-MB-6401) ^17^. The POC-CCA test was only used to detect *S. mansoni* infection because it has low sensitivity for the detection of *S. haematobium* infection ^14,18^. Four visual scores (Negative, 1+, 2+, 3+) were used to provide semi-quantitative results. POC-CCA cassettes were visually read by two technicians, and if the readings were different, the lower reading was taken. They were then read by a cassette reader to estimate the infection intensity, expressed in millivolts (mV), the latter being proportional to the number of worms present. The cutoff of detection was set to 40 mV ^17^.

### Statistical Analysis

Descriptive statistics were used to characterize the study population and to compare urban and rural areas. Chi-square and Student’s *t*-test were used to compare proportions for sex and age between rural and urban children.

The overall prevalence of schistosome infection was calculated based on the number of positive cases of either *S. haematobium* or *S. mansoni*, divided by the total number of tested participants. Since only urine sediment samples negative for *S. haematobium* were examined for *S. mansoni* by POC-CCA, *S. mansoni* prevalence was calculated as the percentage of POC-CCA-positive samples among urine sediment-negative samples. In contrast, *S. haematobium* prevalence was calculated as the percentage of urine sediment samples that were positive among the total number of tested participants. We used the Chi-squared test to compare the proportion of positive schistosomiasis cases, and the Wilcoxon rank-sum test for the mean of infection intensity and egg burden between rural and urban, age group, and sex.

Logistic regression was performed for urban and rural areas, respectively, to determine their similar exposure (sex, age, activities in the river, water origin, and the distance between schools and water bodies) to schistosome infection. Distance between schools and rivers was calculated with GPS Visualizer using GPS coordinates collected during the survey ^19^. Linear regression was used to determine the association between exposure and the infection intensity determined by the POC-CCA-Reader with log-transformed values (log(X+1)) and Urine sedimentation (eggs per 10 mL) separately. R Version 4.2.1 was used for statistical analysis and visualization.

### Ethics considerations

This study was approved by the National Ethics Committee of the Ministry of Public Health in Kinshasa, Democratic Republic of the Congo (Approval reference number 1254/CLNGA/PCE/PT/1138/NK/2018). School directors met with parents to inform about the study details (purpose, timing, methods and outcome). Those who did not want their children to be enrolled, were allowed to keep them home. According to the Health District Office policy regarding school investigation, school directors were allowed to give signed consent on behalf of the parents. Children gave verbal assent before urine sample collection. Samples and data were anonymously recorded and assigned a code for processing and analysis. Physical addresses were collected from children for follow-up with the treatment at home, also after parents’ consent. Drugs were provided by the NTD program office in Matadi based on the prevalence found.

## Results

### Sociodemographic characteristics of the study population

A total of 1699 schoolchildren who met the inclusion criteria (between 5 and 15 years old, treated at least 6 months before the survey) were included in the study. They were recruited from 10 schools, four (Kivuvu II, Kivuvu III, Nsunda Gallant, Bamanku) from rural and six (Malanga I, Malanga II, Kiasungua I, Kiasungua II, Wene, Kilueka) from urban areas (Fig. 2). Schoolchildren from the rural areas were slightly older than those from urban areas, with a mean age of 10.4 years and 10.1 years, respectively, but this difference was not statistically significant (t = 1.797; p = 0.07). In both areas, there was no significant difference in proportions of boys and girls who participated, with 869 males and 830 females included (χ2 = 3.56; p = 0.34) (Table 1).

**Figure 1.**
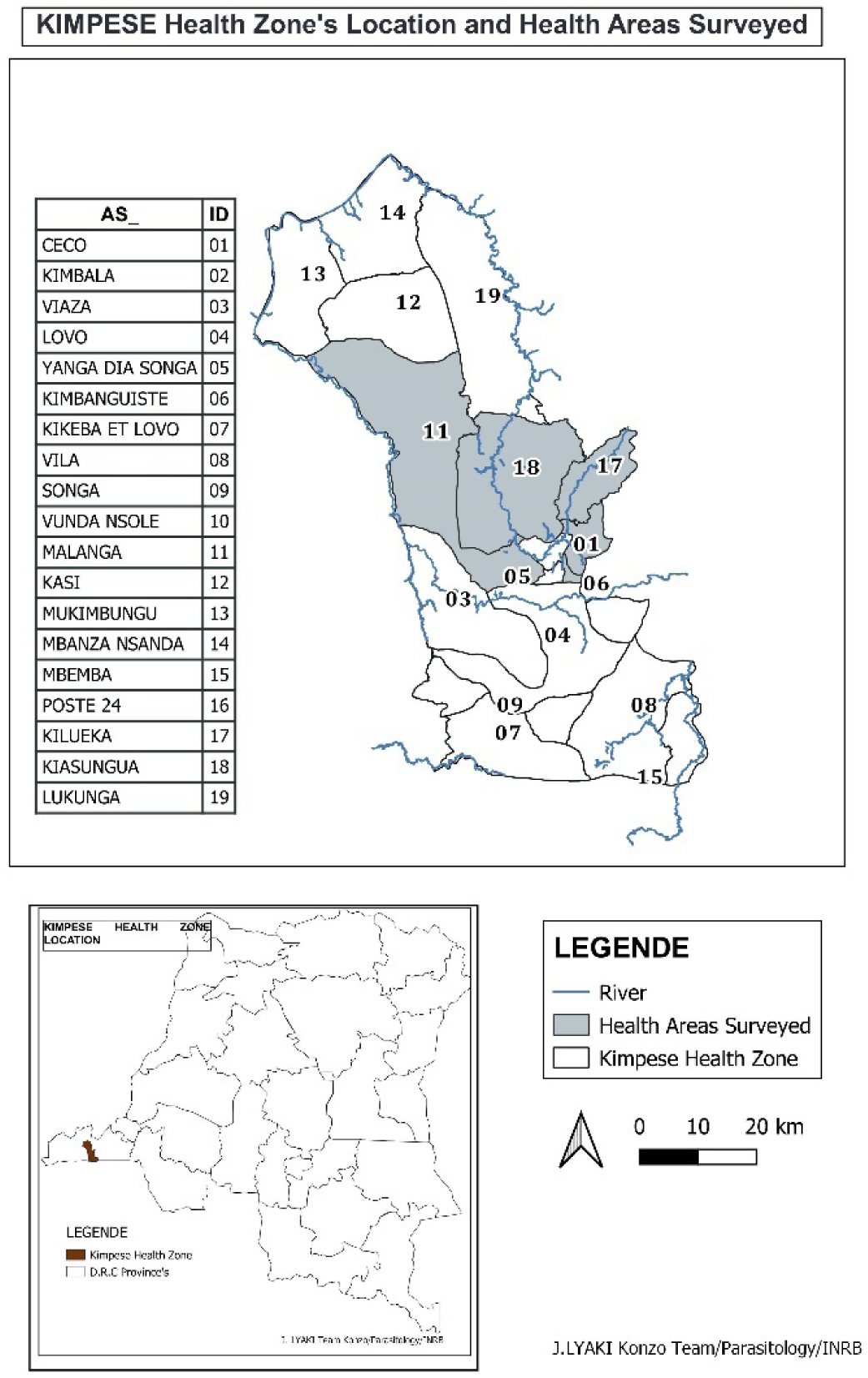
Kimpese health district map showing all 19 health areas with five of them surveyed (in grey)

**Figure 2.**
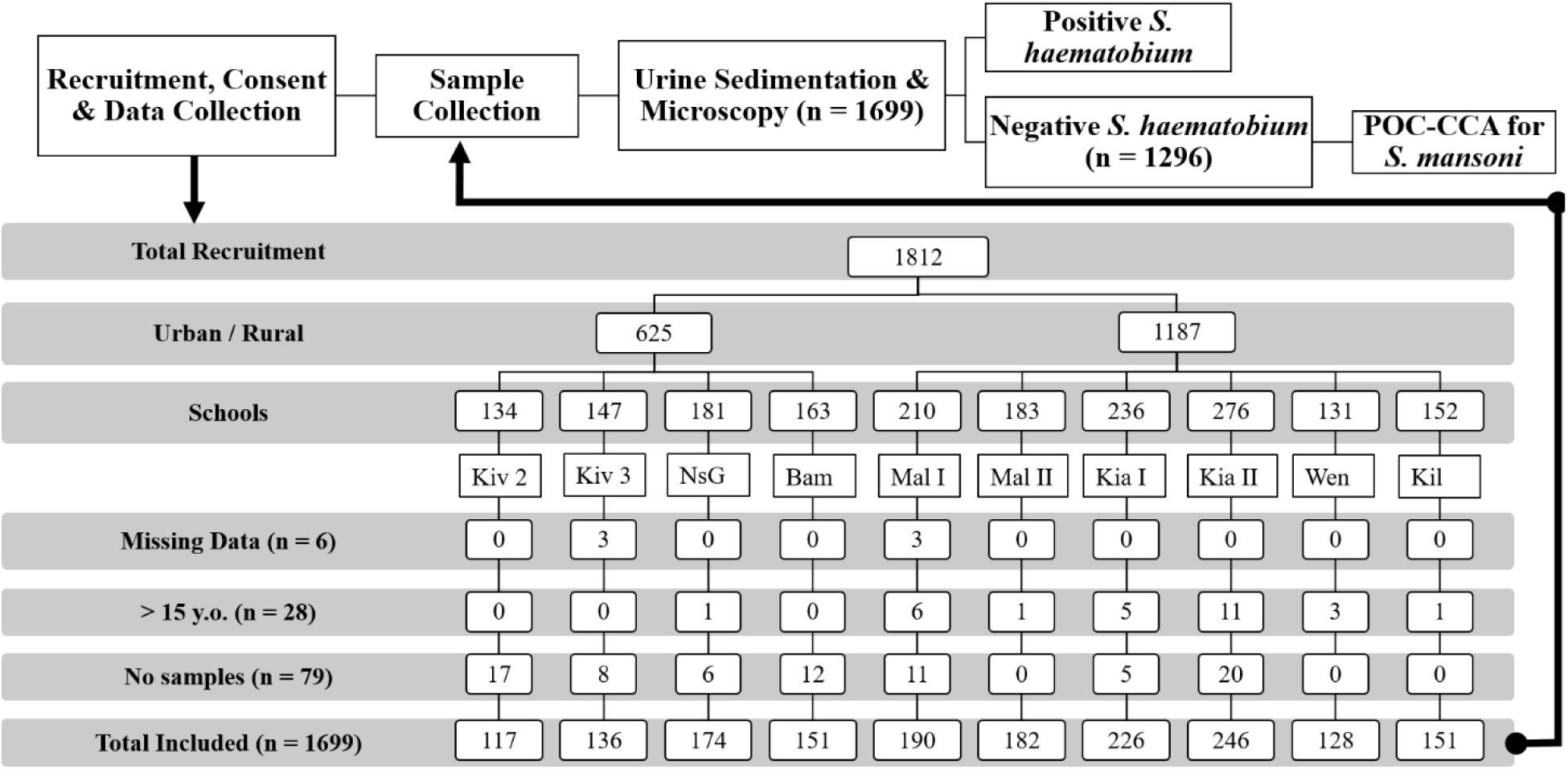
Workflow of the recruitment procedure and data collection and analysis. Kiv 2: Kivuvu II; Kiv 3: Kivuvu III; NsG: Nsunda Gallant; Bam: Bamanku; Mal I: Malanga I; Mal II: Malanga II; Kia I: Kiasungua I; Kia II: Kiasungua II; Wen: Wene; Kil: Kilueka.

**Table 1.** Socio-demographic characteristics of school participants (N = 1699)

| **Socio-demographic profile** | **Number (n = 1699)** | **Frequency (%)** |
| --- | --- | --- |
| **Sex** |  |  |
| **Female** | 830 | 48.9 |
| **Male** | 869 | 51.1 |
| **Age categories** |  |  |
| **5 – 10 years** | 815 | 48 |
| **11 – 15 years** | 884 | 52 |
| **Area Type** |  |  |
| **Urban** | 577 | 34 |
| **Rural** | 1122 | 66 |
| **Health Areas** |  |  |
| **CECo** | 427 | 25.1 |
| **Kimbanguiste** | 150 | 8.9 |
| **Malanga** | 371 | 21.8 |
| **Kiasungua** | 472 | 27.8 |
| **Kilueka** | 279 | 16.4 |
| **River Activities** |  |  |
| **Dishes** | 833 | 49 |
| **Laundry** | 1140 | 67.1 |
| **Swimming** | 899 | 52.9 |
| **Water Origin** |  |  |
| **Well** | 343 | 20.2 |
| **River** | 419 | 24.7 |
| **Spring** | 395 | 23.2 |
| **Tap** | 542 | 31.9 |

### Schistosomiasis prevalence

The overall prevalence of schistosomiasis was 80%, of *S. haematobium* infection 24%, and of *S. mansoni* infection 73%. Schistosomiasis prevalence was higher in the rural than in the urban area (82% vs 75%, X^2^ = 12.22, *p* = .00005). *S. haematobium* infection was more prevalent in the urban area than in the rural area (37% vs 17%, X^2^ = 87.46, *p* < 2.2E^-16^), whereas, in contrast, *S. mansoni* infection was more prevalent (78% vs 59%, X^2^ = 4045, *p* < 2.02E^-10^) (Table 2).

**Table 2.**
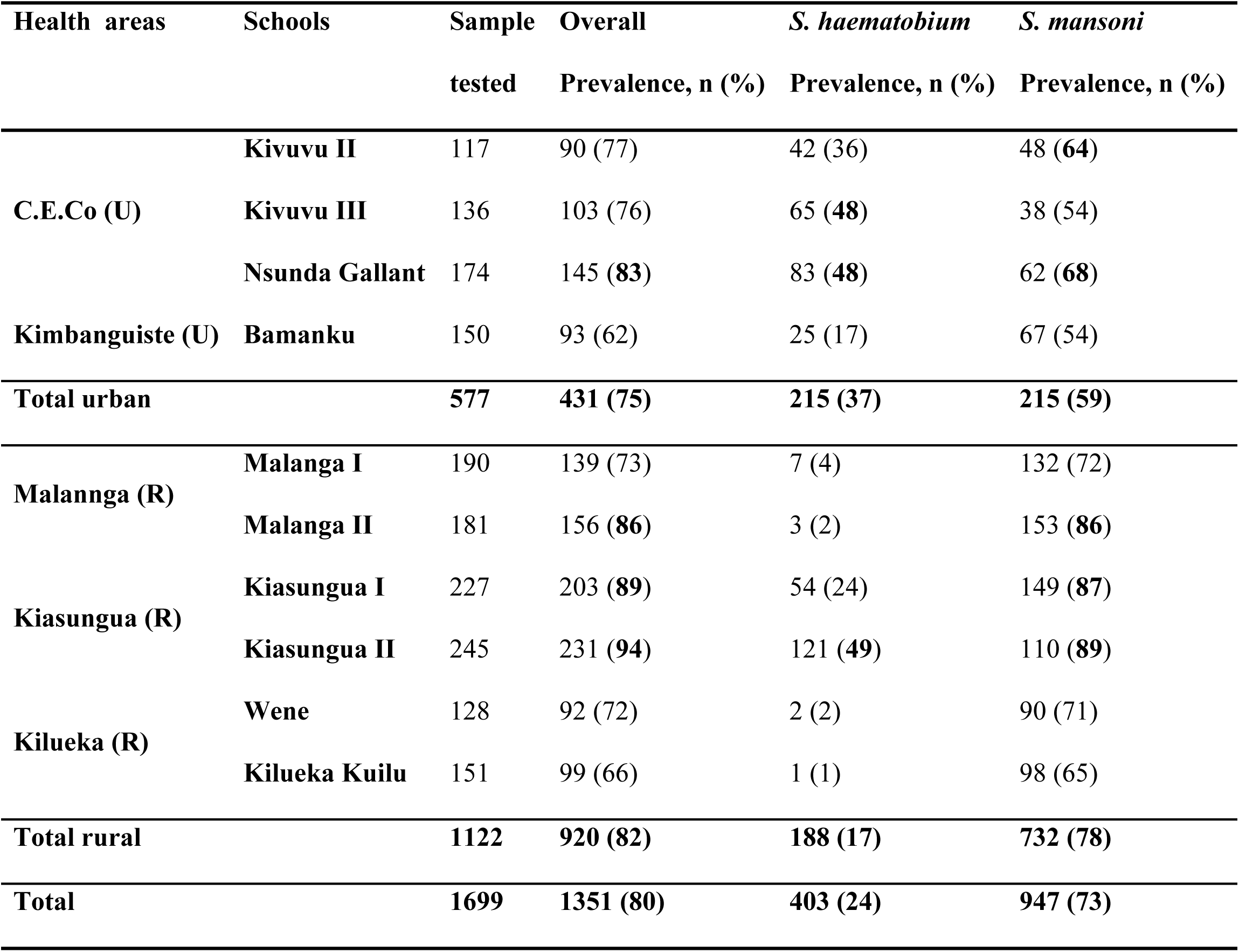
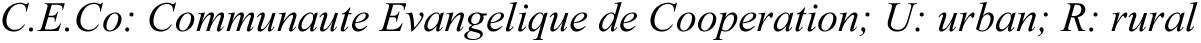
Numbers of cases and prevalences in each school and total. Percentages in bold indicate the highest prevalence.

| Health areas | Schools | Sample tested | Overall Prevalence, n (%) | *S. haematobium* Prevalence, n (%) | *S. mansoni* Prevalence, n (%) |
| --- | --- | --- | --- | --- | --- |
| C.E.Co (U) | Kivuvu II | 117 | 90 (77) | 42 (36) | 48 (**64**) |
|  | Kivuvu III | 136 | 103 (76) | 65 (**48**) | 38 (54) |
|  | Nsunda Gallant | 174 | 145 (**83**) | 83 (**48**) | 62 (**68**) |
| Kimbanguiste (U) | Bamanku | 150 | 93 (62) | 25 (17) | 67 (54) |
| Total urban |  | 577 | 431 (**75**) | 215 (**37**) | 215 (**59**) |
| Malannga (R) | Malanga I | 190 | 139 (73) | 7 (4) | 132 (72) |
|  | Malanga II | 181 | 156 (**86**) | 3 (2) | 153 (**86**) |
| Kiasungua (R) | Kiasungua I | 227 | 203 (**89**) | 54 (24) | 149 (**87**) |
|  | Kiasungua II | 245 | 231 (**94**) | 121 (**49**) | 110 (**89**) |
| Kilueka (R) | Wene | 128 | 92 (72) | 2 (2) | 90 (71) |
|  | Kilueka Kuilu | 151 | 99 (66) | 1 (1) | 98 (65) |
| Total rural |  | 1122 | 920 (**82**) | 188 (17) | 732 (**78**) |
| Total |  | 1699 | 1351 (**80**) | 403 (24) | 947 (73) |

C.E.Co: Communauté Evangelique de Cooperation; U: urban; R: rural

School-wise, Kiasungua II, Kiasungua I, Malanga II, and Nsunda Gallant had higher overall schistosomiasis prevalence of at least 80%. Concerning *S. haematobium* prevalence, it was higher in Kiasungua II, Nsunda Gallant, and Kivuvu III (> 40%), and for *S. mansoni*, Kiasungua II, Kiasungua I, Malanga II, Nsunda Gallant, and Kivuvu II had higher prevalence (> 60%) (Table 2).

### Intensity of S. haematobium & S. mansoni infection

Children in rural areas had higher intensity of infection than those in urban areas for both *S. mansoni* (W = 212056, p < 2.2E^-16^; CI95%: 30.04 to 48.29) and *S. haematobium* (W = 261961, p < 2.2E^-16^; CI95%: -5.62E^-5^ to -2.46E^-5^) (Fig 3A & B). Kiasungua, in the rural area, and CECo, in the urban area, had the highest *S. mansoni* and *S. haematobium* infection intensity (Fig 3C & D). *S. mansoni* infection intensity was higher in older children when compared to younger children (p < 0.001) only in the rural area, whereas *S. haematobium* infection intensity was higher in older children in both rural and urban areas (p < 0.05) (Fig 3E & F). Although the trend was toward higher mansoni schistosomiasis intensity in boys in both rural and urban areas, there were no statistical differences (Fig 3G). Only boys showed significantly higher egg burden for haematobium schistosomiasis in the rural area (p < 0.01) (Fig 3H).

**Figure 3.**
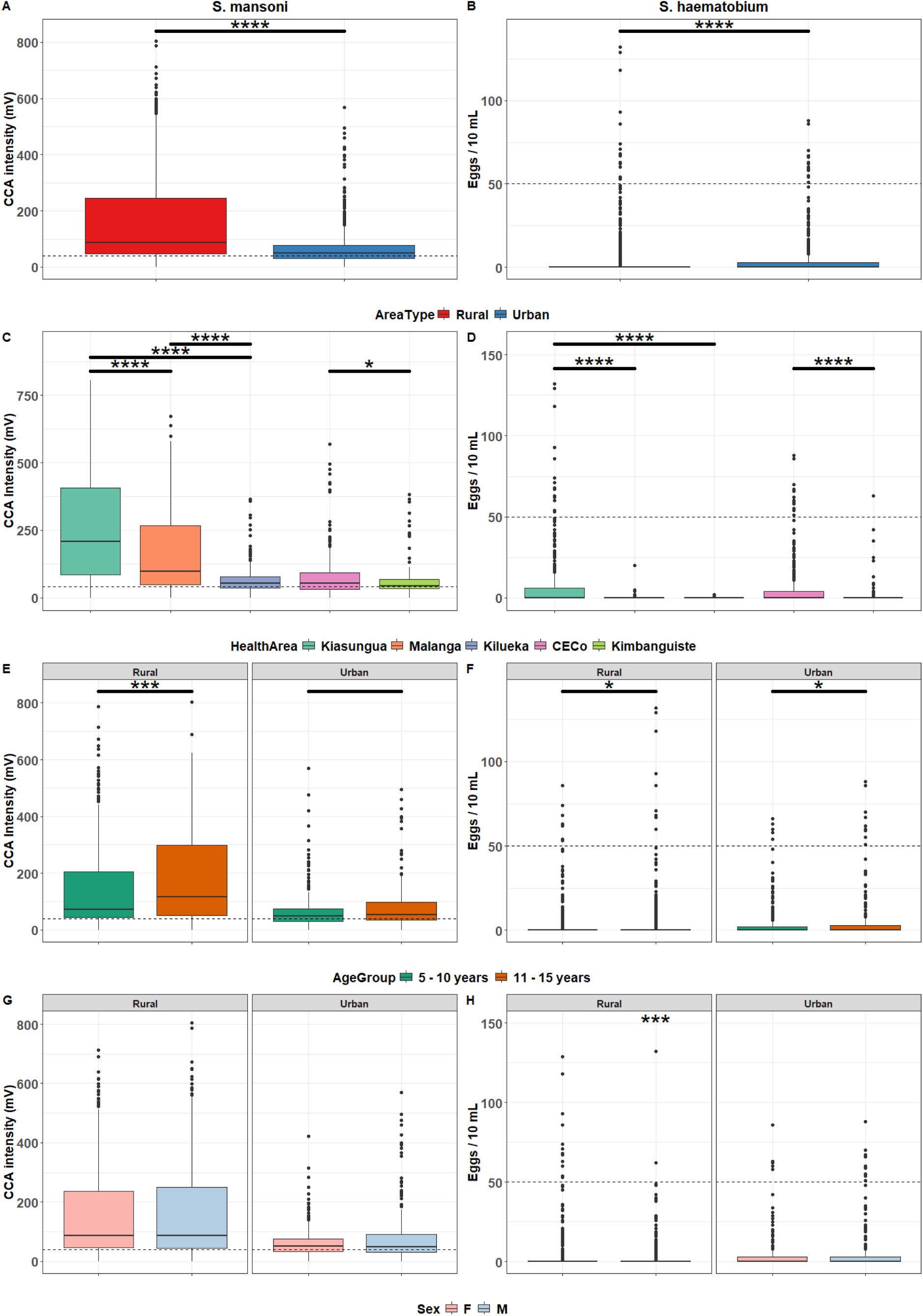
Intensity of *S. mansoni* (left plots) and *S. haematobium* (right plots) infection measured with the POC-CCA and urine sedimentation, respectively, in rural versus urban areas (Area types, health areas, age groups, and sex). In *S. mansoni* plots, the dashed line represents the positivity threshold set for the POC-CCA (40 mV), while in the *S. haematobium* plots it shows the heavy egg burden threshold (50 eggs / 10 mL).

### Factors associated with prevalence and intensity of schistosomiasis infection

Children were asked about the source of the water used at home and whether they were regularly involved in specific activities in the river. The source of water available for domestic purposes varied substantially between health districts. Children from the urban site mainly got water from the tap. In the rural site, the majority of those from Kiasungua used spring water, while most of the children from Kilueka were in contact with the river and those from Malanga usually got their water from wells (Table 3).

**Table 3.** Factors associated with *S. mansoni* schistosomiasis intensity of infection in urban and rural areas. Significant p-values and odds ratios are shown in bold.

| Schistosomiasis prevalence |  |  |  |  | *S. mansoni* intensity |  |  |  | *S. haematobium* intensity |  |  |  |
| --- | --- | --- | --- | --- | --- | --- | --- | --- | --- | --- | --- | --- |
| Urban |  |  | Rural |  | Urban |  | Rural |  | Urban |  | Rural |  |
| Factors | *p* | OR<br>(95% CI) | *p* | OR<br>(95% CI) | *p* | OR<br>(95% CI) | *p* | OR<br>(95% CI) | *p* | OR<br>(95% CI) | *p* | OR<br>(95% CI) |
| Sex (Male) | n.s. | 0.75<br>(0.47 – 1.18) | n.s. | 0.85<br>(0.59 - 1.23) | n.s. | 0.95<br>(0.67 – 1.36) | n.s. | 0.95(0.79 – 1.13) | n.s. | 0.99<br>(0.79 – 1.22) | n.s. | 0.97<br>(0.94 – 1.00) |
| Age | n.s. | 1.08<br>(0.96 – 1.21) | n.s. | 0.99<br>(0.92 - 1.07) | n.s. | 0.97<br>(0.88 – 1. 1,36) | n.s. | 1.03<br>(0.99 – 1.07) | <0.01 | 1.08<br>(1.03 – 1.14) | n.s. | 1.00<br>(0.99 – 1.01) |
| Activities in river |  |  |  |  |  |  |  |  |  |  |  |  |
| Dishes | n.s. | 1.56<br>(0.69 – 3.52) | n.s. | 0.91<br>(0.51 – 1.63) | 0.04 | 1.89<br>(1.01 – 3.51) | n.s. | 1.00<br>(0.78 – 1.28) | n.s. | 0.98<br>(0.69 – 1.41) | n.s. | 0.98<br>(0.93 – 1.02) |
| Laundry | n.s. | 1.23<br>(0.66 – 2.32) | 0.02 | 2.17<br>(1.09 – 4.29) | n.s. | 0.85<br>(0.51 – 1.42) | 0.03 | 1.47<br>(1.03 – 1.12) | n.s. | 1.19<br>(0.90 – 1.59) | n.s. | 1.05<br>(0.99 – 1.12) |
| Swimming | 0.02 | 1.76<br>(1.05 – 2.93) | n.s. | 0.66<br>(0.38 – 1.16) | n.s. | 1.34<br>(0.88 – 2.05) | n.s. | 0.97<br>(0.75 – 1.25) | <0.001 | 1.52<br>(1.20 – 1.91) | n.s. | 1.02<br>(0.98 – 1.06) |
| Water source |  |  |  |  |  |  |  |  |  |  |  |  |
| River | - | - | 0.02 | 0.44<br>(0.22 – 0.89) | - | - | n.s. | 0.75<br>(0.54 – 1.06) | - | - | n.s. | 1.05<br>(0.99 – 1.12) |
| Spring | n.s. | 2.84<br>(0.52 – 15.5) | n.s. | 0.87<br>(0.47 – 1.61) | n.s. | 0.61<br>(0.17 – 2.23) | n.s. | 0.97<br>(0.72 – 1.30) | n.s. | 1.39<br>(0.57 – 3.35) | n.s. | 1.03<br>(0.98 – 1.08) |
| Tap | n.s. | 1.23<br>(0.30 – 5.01) | - | - | n.s. | 0.54<br>(0.18 – 1.65) | - | - | n.s. | 1.14<br>(0.53 – 2.46) |  |  |
| Distance between school and river |  |  |  |  |  |  |  |  |  |  |  |  |
| Linear | - | - | - | - | - | - | - | - | - | - | 0.02 | 0.94 |
|  |  |  |  |  |  |  |  |  |  |  |  | (0.90 – 0.99) |
| Quadratic | <0.001 | 2.45 (1.63 – 3.68) | 0.03 | 0.61 (0.39 – 0.96) | n.s. | 0.74 | <0.001 | 0.66 | <0.0001 | 1.73 | - | - |
|  |  |  |  |  |  | (0.52 – 1.05) |  | (0.53 – 0.80) |  | (1.41 – 2.13) |  |  |

*P*: p-values; OR: Odds Ratio; n.s: non-significant p-values

Logistic regression also showed a very strong association of schistosomiasis infection with the Nsunda Gallant school in the urban area (p = 1.9 × 10^-06^, OR = 3.97 (CI95%: 2.25 - 7.01)).

## Discussion

Our study found an overall schistosomiasis prevalence of 80%, with 73% and 24% for intestinal mansoni schistosomiasis and urinary schistosomiasis, respectively (Table 2), with an average infection intensity of 119 mV by POC-CCA, with age, sex, and water body contact as risk factors associated with *Schistosoma* infection /and its intensity. In 2021, a survey in Kisangi District, which is close to Kimpese District, revealed an overall prevalence of 55%, with 36% for *S. mansoni,* 41% for *S. haematobium,* and 21% for both *S. mansoni* and *S. haematobium* coinfection using, respectively, the Kato-Katz and urinary filtration tests ^6^.

Only one study in Malanga, a rural site in the Kimpese health district, showed a prevalence of 29% for *Schistosoma spp* using Kato-Katz ^20^, compared to our results showing over twice that prevalence using the POC-CCA (79%).

Overall, the rural children had higher intensity of infection than urban children (p < 2.2e-16). Many factors can explain such a difference. Our survey was held during the dry season with cooler weather and low and more stable water. Risk of infection and reinfection might be greater during this period, but might not have much impact on the prevalence since schistosomiasis is a long-lasting infection. In Ethiopia, season, along with water body characteristics, were significantly associated with the abundance of snails. The *Biomphalaria* snail abundance was higher in colder weather (winter) than in hot weather (summer). The *Biomphalaria* population can also vary across sites and depends on the type of water (lake or river). They also suggested that snails may prefer more stable water conditions for survival, which can increase their abundance, and consequently increase the risk of infection in sites with high human-water contact, and reinfection in areas under MDA ^21,22^.

Whilst there was no overall difference in prevalence of schistosomiasis in urban and rural areas, there were significant differences in the distribution of the two species. *S. haematobium* had higher prevalence in urban areas while *S. mansoni* had higher prevalence in rural areas (Table 2). This can be explained by children’s behavior. In rural places, due to a lack of proper sanitation and hygiene, children tend to defecate in the river and/or in the vegetation around, which would then be washed into the river by the rain, therefore increasing the risk of transmission of *S. mansoni* infection. In urban areas, on the contrary, because of the existence of proper sanitation, children may not defecate in the rivers. However, when being active in the rivers for swimming, for instance, and being far from the sanitaries, they may tend to urinate in the river, increasing in the same way the risk of transmitting *S. haematobium* infection.

Age had a small but significant effect on infection intensity in rural areas (OR = 1.1; p = 0.002) for *S. haematobium* infection. Specifically, children from 11 to 15 years old showed significantly higher intensity of infection (Fig 3F) in both rural and urban area settings. In Cameroon, a field assessment of the POC-CCA cassette reader for the quantification of *S. mansoni* infection intensity showed a significant association with age (5 - 14 years old) in school children (p = 0.00022) ^17^. Although age is not associated with S. mansoni infection for the schoolchildren of the Kimpese health zone, those from 11 to 15 years old in the rural area showed a very high intensity of infection compared to those from 5 to 10 years old. This difference can be explained by the fact that, considering the distance between schools and rivers (Table S1), older children are more likely to walk relatively long distances after school to get to those rivers than younger ones.

The endemicity of both mansoni and haematobium schistosomiasis in an area increases the risk of dual infections. There are significant proportions of dual infections in endemic areas in Ghana and Tanzania, as well as in other foci throughout DRC ^23,24^. We did not assess co-infections in our survey as we used POC-CCA to only detect *S. mansoni* infection among *S. haematobium* negative participants. However, although POC-CCA is more sensitive to *S. mansoni*, it can also detect *S. haematobium* infection with low sensitivity, but similar specificity for *S. mansoni* detection ^13^. It is, therefore, possible that among *S. mansoni* positive individuals are also *S. haematobium* positive participants with very low worm burden or no egg excretion. Additionally, the evidence from studies of mixed infections is inconclusive; in Ghana the prevalence of *S. mansoni* schistosomiasis was similar in those with and without concurrent *S. haematobium* schistosomiasis infections ^23^, whereas in Tanzania there was an excess of *S. mansoni* among those with a concurrent *S. haematobium* infection ^24^. In our study, the prevalence of *S. mansoni* schistosomiasis in Kilueka and Malanga, which had very low *S. haematobium* prevalence, was within the range of values seen in districts with higher *S. haematobium* prevalence (60% - 90%). This was consistent with our *S. mansoni* prevalence not being biased by excluding *S. haematobium*-infected samples.

Multiple regression analysis on children’s behavior activities in the river (laundry, dishes, swimming, etc.) and environmental risk factors (water source) showed significant associations with schistosomiasis infection and its burden. Age and swimming were associated with prevalence, and *S. haematobium* intensity in the Kimpese urban area. Due to their very close proximity to contaminated rivers (less than 0.25km), Kivuvu and Nsunda Gallant schools played a considerable role in schistosomiasis infection. In Uganda, a study found that a proximity of less than 30 minutes away from contaminated water increased the risk of schistosomiasis infection ^25^. In contrast, despite being located over a kilometre away from the water bodies, rural schools in Kiasungua still had high *S. haematobium* egg burden and high intensity of *S. mansoni* infection. This is obvious as those water sources (spring and river) are the only available and closest in the area (Table S1).

Our study documents for the first time the use of the POC-CCA cassette reader for schistosomiasis infection quantification on freshly collected urine samples, contrary to previous studies done on banked samples for areas in the DRC ^26^. The schistosomiasis mansoni investigation was limited to assessing using only the POC-CCA, which is not yet a standard test for that specific infection despite its high sensitivity compared to the Kato-Katz test. Not evaluating coinfection for Schistosoma mansoni and Schistosoma haematobium infections resulted in an estimation of both infection prevalences.

On the other hand, considering its sensitivity, we could detect more infected cases, which could be the same for the other studies that used only Kato-Katz and present a more realistic epidemiologic state of Schistosomiasis in the studied site. Also, its malleability allowed a large-scale and more sensitive screening of intestinal schistosomiasis in a relatively short time.

## Conclusion

This survey revealed differences in the prevalence and the intensity of infection between rural and urban areas, and between health areas within them. The overall difference between urban and rural areas was only moderate, indicating that urban areas should not be overlooked in treatment programs.

There were differences in the prevalence of *S. mansoni* between rural and urban areas and evidence of high *S. haematobium* prevalence in the urban area. *Schistosoma* host snails’ distribution along with environmental parameter investigations could give more insights and a better view on the schistosomiasis exposure risks in these endemic locations. Children’s behavior and contact with snail-infested rivers still determine the prevalence of the infection independently of the presence of secure water sources in the area.

## Data Availability

All relevant data underlying the findings of this study are included within the manuscript and its Supporting Information files. The de-identified dataset used for the analyses is provided as Supporting Information.

## Acknowledgements

The authors thank all the participants for contributing to this study.

Paul Corstjens & Govert Van Dam from Leiden University (Netherlands), developers of the C.C.A. and the C.A.A. tests.

The Evangelic Medical Institute (Kimpese, Kongo Central) for lab availability.

This study was funded by Wellcome Trust through the Science for Africa Foundation grant, [H3A-18-004] to the TrypanoGEN+ group as part of the H3Africa consortium. The views expressed herein are those of the author(s) and do not necessarily reflect those of the SFA Foundation and its partners.

## Author Contributions

### Conceptualization

Kevin Lubula Karume, Harry Noyes, Oscar Nyangiri, Dieudonné Mumba Ngoyi.

### Data curation

Kevin Lubula Karume, Olivier Fataki, Ronald Kyong Shin, Harry Noyes, Julius Mulindwa, Jacent Nassuuna,.

### Formal analysis

Kevin Lubula Karume, Olivier Fataki, Harry Noyes, Oscar Nyangiri, Jacent Nassuuna, Allisson Elliott, Jacob Lyaki.

### Funding acquisition

Dieudonné Mumba Ngoyi.

### Methodology

Kevin Lubula Karume, Oscar Nyangiri, Harry Noyes, Julius Mulindwa, Miriam Casacuberta Partal.

### Project administration

Dieudonné Mumba Ngoyi.

### Supervision

Oscar Kwabanzawoko, Théophile Mbemba, Dieudonné Mumba Ngoyi.

### Writing – original draft

Kevin Lubula Karume, Harry Noyes, Jean Pierre Kambala Mukendi.

### Writing – review & editing

Kevin Lubula Karume, Harry Noyes, Oscar Nyangiri, Jacent Nassuuna, Allisson Elliott, Julius Mulindwa, Jean Pierre Kambala Mukendi.

## Notes

### Competing Interest Statement

The authors have declared no competing interest.

### Author Declarations

The study was approved by the Ethics Committee of the Ngaliema Clinics, Ministry of Public Health, Kinshasa-Gombe (Approval Ref. No. COMETH/TKK/PRES/004/2018). Written informed consent was obtained from the parents or legal guardians of all participating children, and assent was obtained from the children when appropriate.

## References

1. Hailegebriel T, Nibret E, Munshea A. Prevalence of Schistosoma mansoni and S. haematobium in Snail Intermediate Hosts in Africa: A Systematic Review and Meta-analysis. J Trop Med. Hindawi Limited. 2020;2020. doi:10.1155/2020/8850840

2. Chitsulo L, Engels D, Montresor A, Savioli L. The Global Status of Schistosomiasis and Its Control Europe PMC Funders Group. Vol 77. 2000.

3. Klohe K, Koudou BG, Fenwick A, et al. A systematic literature review of schistosomiasis in urban and peri-urban settings. PLoS Negl Trop Dis. Public Library of Science. 2021;15(2):1–19. doi:10.1371/journal.pntd.0008995

4. Madinga J, Linsuke S, Mpabanzi L, et al. Schistosomiasis in the Democratic Republic of Congo: A literature review. Parasit Vectors. BioMed Central Ltd. 2015;8(1). doi:10.1186/s13071-015-1206-6

5. Linsuke S, Nundu S, Mupoyi S, et al. High Prevalence of Schistosoma mansoni in Six Health Areas of – Kasansa Health Zone, Democratic Republic of the Congo: Short Report. PLoS Negl Trop Dis. 2014;8(12). doi:10.1371/journal.pntd.0003387

6. Linsuke S, Ilombe G, Disonama M, et al. Schistosoma Infection Burden and Risk Factors among School-Aged Children in a Rural Area of the Democratic Republic of the Congo. Trop Med Infect Dis. 2023;8(9). doi:10.3390/tropicalmed8090455

7. Colley DG, King CH, Kittur N, et al. Evaluation, validation, and recognition of the point-of-care circulating cathodic antigen, urine-based assay for mapping schistosoma mansoni infections. American Journal of Tropical Medicine and Hygiene. 2020;103:42–49. doi:10.4269/ajtmh.19-0788

8. Bä Renbold O, Garba A, Colley Id DG, et al. Translating preventive chemotherapy prevalence thresholds for Schistosoma mansoni from the Kato-Katz technique into the point-of-care circulating cathodic antigen diagnostic test. Published online 2018. doi:10.1371/journal.pntd.0006941

9. Okoyo C, Simiyu E, Njenga SM, Mwandawiro C. Comparing the performance of circulating cathodic antigen and Kato-Katz techniques in evaluating Schistosoma mansoni infection in areas with low prevalence in selected counties of Kenya: A cross-sectional study. BMC Public Health. 2018;18(1). doi:10.1186/s12889-018-5414-9

10. Alemu G, Nibret E, Munshea A, et al. Comparative performance of Kato–Katz, POC-CCA and real-time PCR in detecting Schistosoma mansoni infection at different endemicity settings in northwest Ethiopia: a cross-sectional study. Tropical Medicine and Health 2025 53:1. 2025;53(1):103-. doi:10.1186/S41182-025-00777-7

11. Isaiah PM, Nyawanda B, Okoyo C, Steinmann P. Kato-Katz versus urine POC-CCA for the diagnosis of Schistosoma mansoni in preschool-aged children in Homa Bay County, Kenya. Parasitol Res. 2025;124(2):25. doi:10.1007/S00436-025-08467-3

12. Colley DG, Binder S, Campbell C, et al. A five-country evaluation of a point-of-care circulating cathodic antigen urine assay for the prevalence of Schistosoma mansoni. American Journal of Tropical Medicine and Hygiene. 2013;88(3):426–432. doi:10.4269/ajtmh.12-0639

13. Lorenz E, Razafindrakoto R, Rausche P, et al. Detecting Schistosoma infections in endemic countries: a diagnostic accuracy study in rural Madagascar. Infect Dis Poverty. 2025;14(1). doi:10.1186/s40249-025-01292-x

14. Sanneh B, Joof E, Sanyang AM, et al. Field evaluation of a schistosome circulating cathodic antigen rapid test kit at point-of-care for mapping of schistosomiasis endemic districts in The Gambia. PLoS One. 2017;12(8). doi:10.1371/journal.pone.0182003

15. Muke Modeste, Konga Moise, Kalonda Christelle. Contribution àl’analyse de l’évolution spatio-temporelle de l’aglomération urbaine de Kimpese (Kongo Central, RD. Congo), entre1979 et 2020, par les techniques géomatiques. Cinq Continents. 2023;13:148–163.

16. Llen A, Oss GPR, Artley ABB, et al. S CHISTOSOMIASIS. Vol 346. 2002. www.nejm.org

17. Mewamba EM, Tiofack AAZ, Kamdem CN, et al. Field assessment in cameroon of a reader of poc-cca lateral flow strips for the quantification of schistosoma mansoni circulating cathodic antigen in urine. PLoS Negl Trop Dis. 2021;15(7). doi:10.1371/journal.pntd.0009569

18. Ayele B, Erko B, Legesse M, Hailu A, Medhin G. Evaluation of circulating cathodic antigen (CCA) strip for diagnosis of urinary schistosomiasis in Hassoba school children, Afar, Ethiopia. Parasite. 2008;15(1):69–75. doi:10.1051/parasite/2008151069

19. GPS Visualizer: Calculators: Great Circle Distance Maps, Airport Routes, & Degrees/Minutes/Seconds. Accessed December 12, 2025. https://www.gpsvisualizer.com/calculators

20. Madinga J, Polman K, Kanobana K, et al. Epidemiology of polyparasitism with Taenia solium, schistosomes and soil-transmitted helminths in the co-endemic village of Malanga, Democratic Republic of Congo. Acta Trop. 2017;171:186–193. doi:10.1016/j.actatropica.2017.03.019

21. Hailegebriel T, Nibret E, Munshea A. Distribution and seasonal abundance of Biomphalaria snails and their infection status with Schistosoma mansoni in and around Lake Tana, northwest Ethiopia. Sci Rep. 2022;12(1). doi:10.1038/s41598-022-21306-0

22. Sokouri EA, Ahouty Ahouty B, N’Djetchi M, et al. Impact of environmental factors on Biomphalaria pfeifferi vector capacity leading to human infection by Schistosoma mansoni in two regions of western Côte d’Ivoire. Parasit Vectors. 2024;17(1). doi:10.1186/s13071-024-06163-2

23. Anyan WK, Pulkkila BR, Dyra CE, et al. Assessment of dual schistosome infection prevalence from urine in an endemic community of Ghana by molecular diagnostic approach. Parasite Epidemiol Control. 2020;9. doi:10.1016/j.parepi.2019.e00130

24. Miller K, Choudry J, Mahmoud ES, Lodh N. Accurate Diagnosis of Schistosoma mansoni and S. haematobium from Filtered Urine Samples Collected in Tanzania, Africa. Pathogens. 2024;13(1). doi:10.3390/pathogens13010059

25. Kim ES, Adriko M, Oseku KC, Lokure D, Webb EL, Sabapathy K. Factors associated with hookworm and Schistosoma mansoni infections among school-aged children in Mayuge district, Uganda. BMC Public Health. 2024;24(1). doi:10.1186/s12889-024-19092-7

26. Hoekstra PT, Madinga J, Lutumba P, et al. Diagnosis of Schistosomiasis without a Microscope: Evaluating Circulating Antigen (CCA, CAA) and DNA Detection Methods on Banked Samples of a Community-Based Survey from DR Congo. Trop Med Infect Dis. 2022;7(10). doi:10.3390/tropicalmed7100315

